# Assessing the diagnostic accuracy of artificial intelligence in detecting cervical pre-cancer from pap smear images

**DOI:** 10.1101/2025.03.09.25323636

**Authors:** Sang’udi Sang’udi, David Muhunzi, Whitefrank Frank, Deogratias Mzurikwao, Jackline Ngowi, Davis Amani, Leah F Mnango, Angela Mlole, Belinda J Njiro, Deogratias Kilasara, Mboka Jacob, Bruno Sunguya

## Abstract

The global burden of cervical cancer, with a notable prevalence in regions like Tanzania, highlights the critical need for timely and accurate diagnosis. The scarcity of pathologists in such areas underscores the importance of developing automated tools for the cytopathological analysis of cervical cancer images to improve diagnostic efficiency and accuracy.

This study investigated the performance of advanced artificial intelligence (AI) algorithms for screening cervical cancer using Pap smear cytological slides from the Centre for Recognition and Inspection of Cells CRIC dataset. Deep learning models were trained on images of both cervical cancer and normal cervix cells, with the evaluation of model performance focusing on specificity, sensitivity, and accuracy.

Among the evaluated convolutional neural network (CNN) architectures—EfficientNetB7, MobileNet, ResNet50, ResNet152, and InceptionNet-V3—EfficientNetB7 emerged as the top performer, demonstrating impressive accuracy, sensitivity and specificity metrics (accuracy: 0.95, sensitivity: 0.84, specificity: 0.97). In contrast, InceptionNet-V3 showed the lowest performance across similar metrics (accuracy: 0.78, sensitivity: 0.35, specificity: 0.87). The study also highlighted the challenges in distinguishing between specific cell classes, particularly between ASC-US and LSIL, due to the subtle nuances of cytomorphological criteria.

The findings suggest that while AI can significantly aid in cervical cancer screening, the complexity of cell image classification demands further exploration, possibly incorporating whole smear analysis or additional contextual information to improve accuracy. Despite the observed classification challenges, such as between ASC-US and LSIL classes, the potential for AI in supporting clinical decision-making in cervical cancer management is evident.

## Introduction

Cervical cancer is preventable and treatable if identified early and given appropriate treatment [1]. Nonetheless, it is the most common cancer in women in 25 countries globally, many of which are in sub-Saharan Africa. Cervical cancer presents a significant global health challenge, with nearly 661,044 new cases and 348,186 deaths worldwide in 2022, making it the eighth-highest cause of cancer death among women [2]. The disease has remained an important indicator of global health inequality with more than 85% of deaths occurring in developing regions and over 90% of the highest incidence rates of cervical cancer occurring in sub-Saharan Africa [3,4]. The estimated age-standardized rate of cervical cancer mortality in East Africa is higher than that of any region in the world [5]. In Tanzania, cervical cancer is the leading cause of female cancers with over 10,241 new cases accounting for 40.7% of all female cancers and 6,525 deaths accounting for 24.2% of all cancer deaths in women in 2020 [6]. The disparity in the burden of cervical cancer between resource-rich and resource-limited countries is primarily attributed to low disease awareness, prevention measures and difficulty in implementing screening programs [7].

With advancements in AI technology, numerous systems employing computational algorithms for automatic cell image analysis have been developed to enhance screening precision and efficiency. Researchers have applied classic machine learning strategies, including support vector machines and manually designed features, for cell segmentation and classification [8]. In recent times, Deep Learning approaches have surpassed former leading machine learning methods in various challenges, such as image classification. Convolutional Neural Network architectures such as ResNet-152, ResNet-50, EfficientNet, MobileNet, and Google’s Inception V3 highlight their pivotal role in enhancing medical image analysis for pre-cancerous cervical lesion classification. ResNet models address the vanishing gradient problem, enabling deeper network training for higher accuracy [9]. EfficientNet offers a balanced scaling method for efficiency and accuracy [10], while MobileNet’s lightweight design enables portable diagnostic applications [11]. Inception V3 excels in complex feature extraction for improved screening accuracy [12]. These architectures collectively advance diagnostic capabilities, aiding early detection and treatment in cervical cancer management.

The standard screening test in resource-limited settings is visual inspection with acetic acid (VIA) as mid-level providers can perform this and allow for immediate treatment. However, the result of VIA is a subjective interpretation resulting in variable performances, and the utility is questionable in resource-limited settings when the number of screening rounds per woman’s lifetime is low [5]. To date, cervical cancer prevention efforts worldwide have focused on screening sexually active women using Pap smears and treating precancerous lesions. Pap smear screening has improved the control of cervical cancer in resource-rich countries. However, Pap smear image analysis when done manually by a pathologist, is a tedious, error-prone, and time-consuming process [13]. Given the significantly imbalanced pathologist-to-patient ratio in resource-limited settings where this research is based, this task becomes even more daunting making it difficult to replicate it in resource-limited settings such as Tanzania. With the burden of cervical cancer projected to rise to 700,000 cases and 400,000 deaths by 2030, the vast majority of this increase being in resource-limited settings, it is imperative to develop automatic screening tools that can detect pre-cancerous lesions in Pap smears [1]. These can be used by medical doctors and other mid-level healthcare providers in primary and secondary health facilities without the need for pathologists as a result improving clinical efficiency and efficacy.

Addressing this gap is critical, as the global burden of cervical cancer is expected to rise, with a significant majority of this increase occurring in resource-limited settings. By focusing on the validation of AI-based screening tools that can be used by medical doctors and mid-level healthcare providers without the need for pathologist oversight, this research aims to offer a scalable, efficient solution to cervical cancer screening that could significantly impact early detection and treatment efforts, thereby reducing the global health inequity associated with this disease.

## Materials and methods

### Study Design and Setting

This experimental study adopted a quantitative approach to evaluate the performance metrics of the selected state-of-the-art AI algorithms in detecting pre-cancerous cervical lesions from Pap smear images. The intervention involved the development of AI algorithms using Convolutional Neural Networks (CNN) for the classification of Pap smear images. The selected CNN architectures for this study included EfficientNetB7, MobileNet, ResNet50, ResNet152, and InceptionNet-V3. The study was conducted at Muhimbili University of Health and Allied Sciences, utilizing a computer equipped with an Intel Core (TM) i7-1165G7 processor, GeForce GTX 1650 Ti GPU, 2.8 GHz (8 CPU), and 16 GB of RAM, running on a Windows 64-bit OS. The Python programming language, version 3.11.4, and the Tensorflow/Keras framework were employed.

### Dataset

The study utilized Pap smear images from the public domain of women aged 21-69 who were screened for cervical cancer. The dataset comprised images in TIF format with dimensions of 1376 × 1020 pixels and a resolution of 150 dpi. The classification followed rigorous protocols by three experts The images were selected through systematic random sampling from the Centre for Recognition and Inspection of Cells (CRIC) Searchable Image Database. The dataset used in this work is part of the cervical cell classification collection. It contains cells from six classes: Normal, Atypical squamous Cell-Undetermined significance (ASC-US), Low-grade Squamous Intraepithelial Lesions (LSIL), Atypical squamous Cell-Cannot exclude High grade (ASC-H), High-grade squamous Intraepithelial Lesions (HSIL), and carcinoma (CA). Data acquisition accessed 15/02/2023 – Downloaded six-class dataset which are found in training, validation, and testing folders.

### Data Division

In our study, we utilized the hold-out method to evaluate model generalizability, which entailed handling new, unseen data using a six-class classification system for cervical lesion detection. The dataset was divided into a training set (80% of the images), with a further 20% of this subset designated for validation to monitor the model’s learning process early on. The remaining 20% constituted the test set, used to assess the model’s ability to generalize. This classification approach allowed for precise reporting and detailed tracking of the types of lesions detected across different experimental phases, as outlined in Table 1.

**Table 1.** The number of images from each class for the training, validation, and testing sets.

| Class | ASCH | ASCUS | CA | HSIL | LSIL | Normal | Total |
| --- | --- | --- | --- | --- | --- | --- | --- |
| Training | 385 | 361 | 331 | 429 | 405 | 435 | 2346 |
| Testing | 238 | 211 | 214 | 265 | 247 | 262 | 1437 |
| Validation | 183 | 170 | 142 | 180 | 166 | 165 | 1006 |
| Total per class | 806 | 742 | 687 | 874 | 818 | 862 | 4789 |

### Data preprocessing

The dataset was preprocessed to prepare the images for feature extraction, essential for accurate screening. Utilizing Python, the images were initially processed to visualize their pre- and post-processing states, ensuring they were suitable for further steps. Preprocessing techniques such as zooming, rotation, shearing, and flipping were employed, followed by image enhancement strategies that improved contrast and readability. The images were standardized in size and converted to grayscale to facilitate the CNN’s feature recognition.

We progressed to model development, where the dataset had been previously divided into training, validation, and testing subsets. The training and validation sets were instrumental in training the model and evaluating its learning process, while the test set assessed its performance. The CNN performed feature extraction at various layers, processing the images to classify them according to the stages and classes of cervical cancer. This step was crucial for ensuring that the transformations applied during preprocessing did not alter the class labels, thus maintaining the integrity of the diagnostic process.

### Data Analysis

We utilized five metrics to assess the models’ performance: precision, recall, F1-score, accuracy, and specificity, based on the values of true positives (TPs), true negatives (TNs), false positives (FPs), and false negatives (FNs). The analysis considered two scenarios: one where the test indicates the presence of a lesion, and another where it indicates absence. In the first scenario, a positive result in the presence of a lesion is considered a true positive, whereas a positive result in the absence of a lesion is deemed a false positive. In the second scenario, a negative result when a lesion is present is a false negative, and a negative result in the absence of a lesion is a true negative. Precision (Prec.), defined by Equation (1), evaluates the test’s ability to correctly identify lesions in patients who have them.

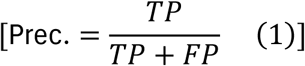

Recall (Rec.), computed using Equation (2), assesses the test’s efficacy in detecting lesions whenever they are present.

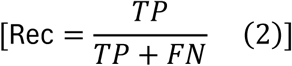

The F1-score, outlined in Equation (3), is the harmonic mean of precision and recall, reflecting the overall efficacy of the proposed methodology.

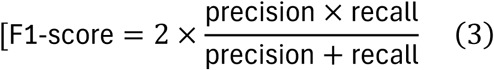

Accuracy (Acc.), indicated by Equation (4), measures the ratio of all correctly identified tests (both positive and negative) against the total number of tests conducted.

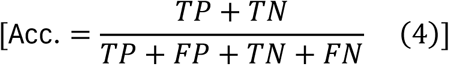

Lastly, specificity (Spec.), shown in Equation (5), evaluates the test’s ability to correctly identify the absence of lesions, i.e., the proportion of individuals without lesions who receive negative test outcomes.

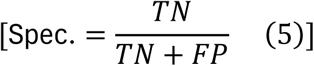

### Ethical Consideration

Ethical clearance was obtained from the MUHAS Institutional Review Board on 23/02/2023 with MUHAS-REC-02-2023-1542 approval number. In the study context, obtaining consent from individuals was not a prerequisite due to the nature of the data collection process, which exclusively involved the use of publicly available online data. This approach ensured that all information utilized in the study had been previously shared in public domains, thereby negating the need for direct consent from individuals. The ethical considerations around data privacy and personal consent were addressed by relying solely on data that has been willingly made accessible to the public by the Center for Recognition and Inspection of Cells (CRIC).

## Results

The total number of images used in this study was 4,789 divided into six classes as explained above. The distribution of cervical cells was almost balanced for each class with HSIL class having the largest number of 874, and ASC-US class having lowest number of 742 marks. This distribution is shown in Figure1

**Fig 1:**
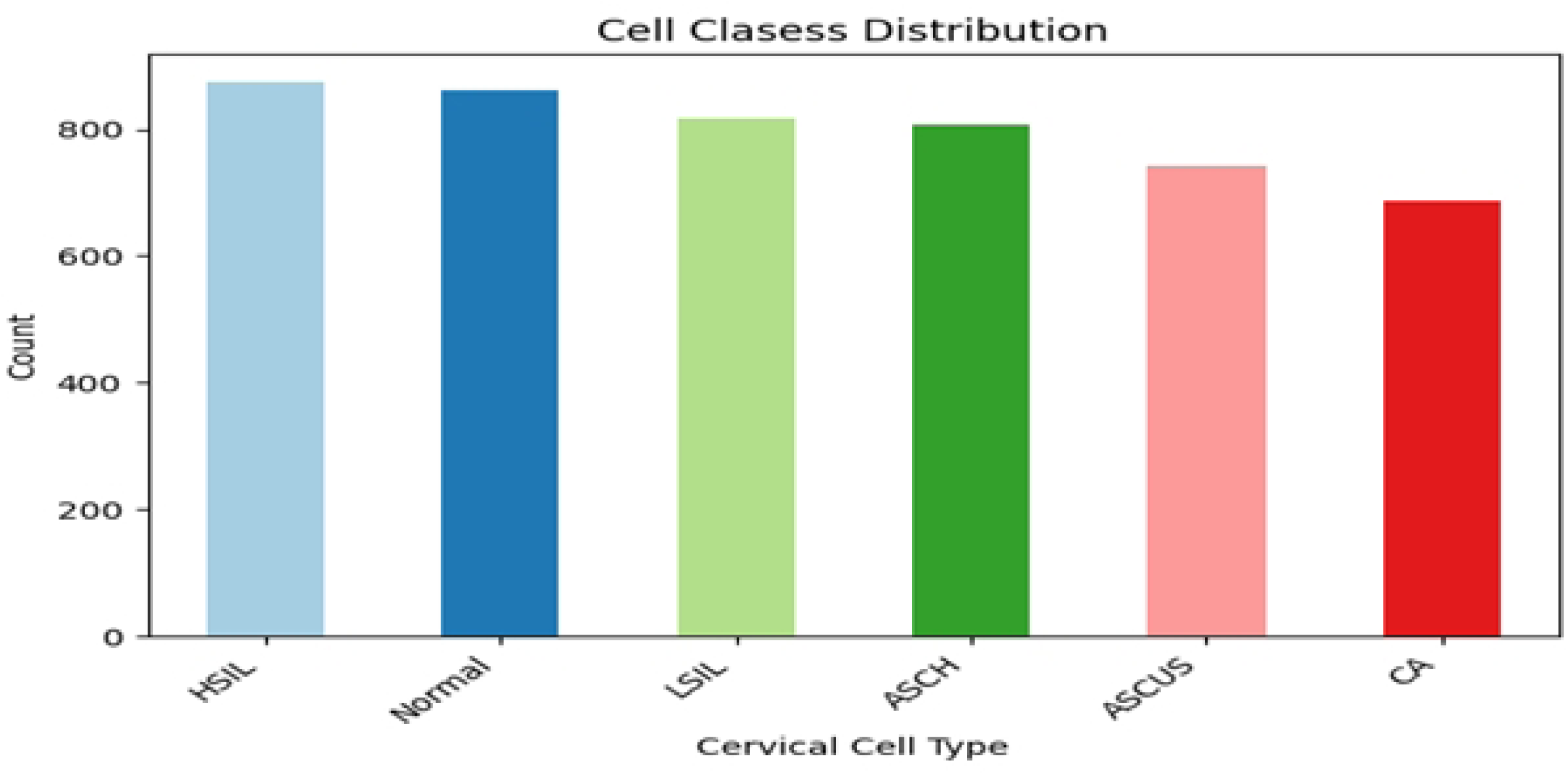
Cervical cells count by each class.

We evaluated the selected algorithms’ performance for cervical cancer screening using Pap smear images. Specifically, we compared the performance metrics of EfficientNetB7, MobileNet, ResNet50, ResNet152, and InceptionNet-V3 in terms of F1-Score, Precision, Recall, Accuracy, Specificity, and Sensitivity. These metrics are crucial for understanding how well each model can identify and classify cervical intraepithelial neoplasia (CIN) from cytological images, which is a key step in early detection and treatment of cervical cancer.

The EfficientNetB7 architecture demonstrated the highest performance across all evaluated metrics. It achieved an F1-Score, Precision, and Recall of 0.84, indicating a strong balance between precision and recall in its predictions. This model also showed an impressive Accuracy of 0.95, along with a Specificity of 0.97 and a Sensitivity of 0.84. The high specificity indicates that EfficientNetB7 is particularly effective at correctly identifying negative cases, while the high sensitivity reflects its ability to correctly identify positive cases.

MobileNet, the second-best performing architecture, exhibited an F1-Score, Precision, and Recall of 0.77, along with an Accuracy of 0.92. It also demonstrated a Specificity of 0.95 and a Sensitivity of 0.77. These results suggest that MobileNet, while not as effective as EfficientNetB7, still performs reasonably well in screening for cervical cancer.

ResNet50 and ResNet152 architectures showed moderate to lower performance compared to EfficientNetB7 and MobileNet. ResNet50 achieved an F1-Score, Precision, and Recall of 0.57, with an Accuracy of 0.86, a Specificity of 0.91, and a Sensitivity of 0.57. ResNet152 had even lower performance, with an F1-Score, Precision, and Recall of 0.5, an Accuracy of 0.83, a Specificity of 0.9, and a Sensitivity of 0.5. These results indicate that while ResNet architectures are capable of detecting lesions in PAP smear images, their performance is significantly outmatched by EfficientNetB7 and MobileNet.

InceptionNet-V3 had the lowest performance among the evaluated models, with an F1-Score, Precision, and Recall of 0.35. Its Accuracy was 0.78, with a Specificity of 0.87 and a Sensitivity of 0.35. This suggests that InceptionNet-V3 is less suitable for the task of cervical cancer screening compared to the other evaluated architectures.

In summary, our results indicate that EfficientNetB7 and MobileNet are the most promising deep learning architectures for the task of automated cervical cancer screening using PAP smear images. Their high accuracy, sensitivity, and specificity highlight their potential for integration into point-of-care (POC) facilities, especially in low and middle-income countries where the burden of cervical cancer is highest and resources for expert analysis are limited. These findings underscore the importance of selecting the appropriate model architecture for developing AI-based tools for cervical cancer screening and early detection.

**Table 2:** Performance metrics for model Training.

| Architecture | F1-Score | Precision | Recall | Accuracy | Specificity | Sensitivity |
| --- | --- | --- | --- | --- | --- | --- |
| EfficientNetB7 | 0.84 | 0.84 | 0.84 | 0.95 | 0.97 | 0.84 |
| MobileNet | 0.77 | 0.77 | 0.77 | 0.92 | 0.95 | 0.77 |
| ResNet50 | 0.57 | 0.57 | 0.57 | 0.86 | 0.91 | 0.57 |
| ResNet152 | 0.5 | 0.5 | 0.5 | 0.83 | 0.9 | 0.5 |
| InceptionNet-V3 | 0.35 | 0.35 | 0.35 | 0.78 | 0.87 | 0.35 |

The confusion matrix generated by EfficientNetB7 for the classification of six different classes indicated a notable confusion between the ASC-US and LSIL classes. A distinguishing characteristic between these classes is the frequency of occurrence within the smear. Cells are classified as LSIL when numerous occurrences are observed, whereas fewer occurrences result in an ASC-US classification. This study, focusing on the analysis of individual cell images, could not adopt this differentiation strategy, as it necessitates the evaluation of an entire smear. Additionally, the methodology does not account for the interrelations among cropped images. Nonetheless, it’s important to note that both ASC-US and LSIL classes result in the same clinical recommendation: a follow-up and re-examination in six months to a year, depending on the patient’s age.

**Fig 2:**
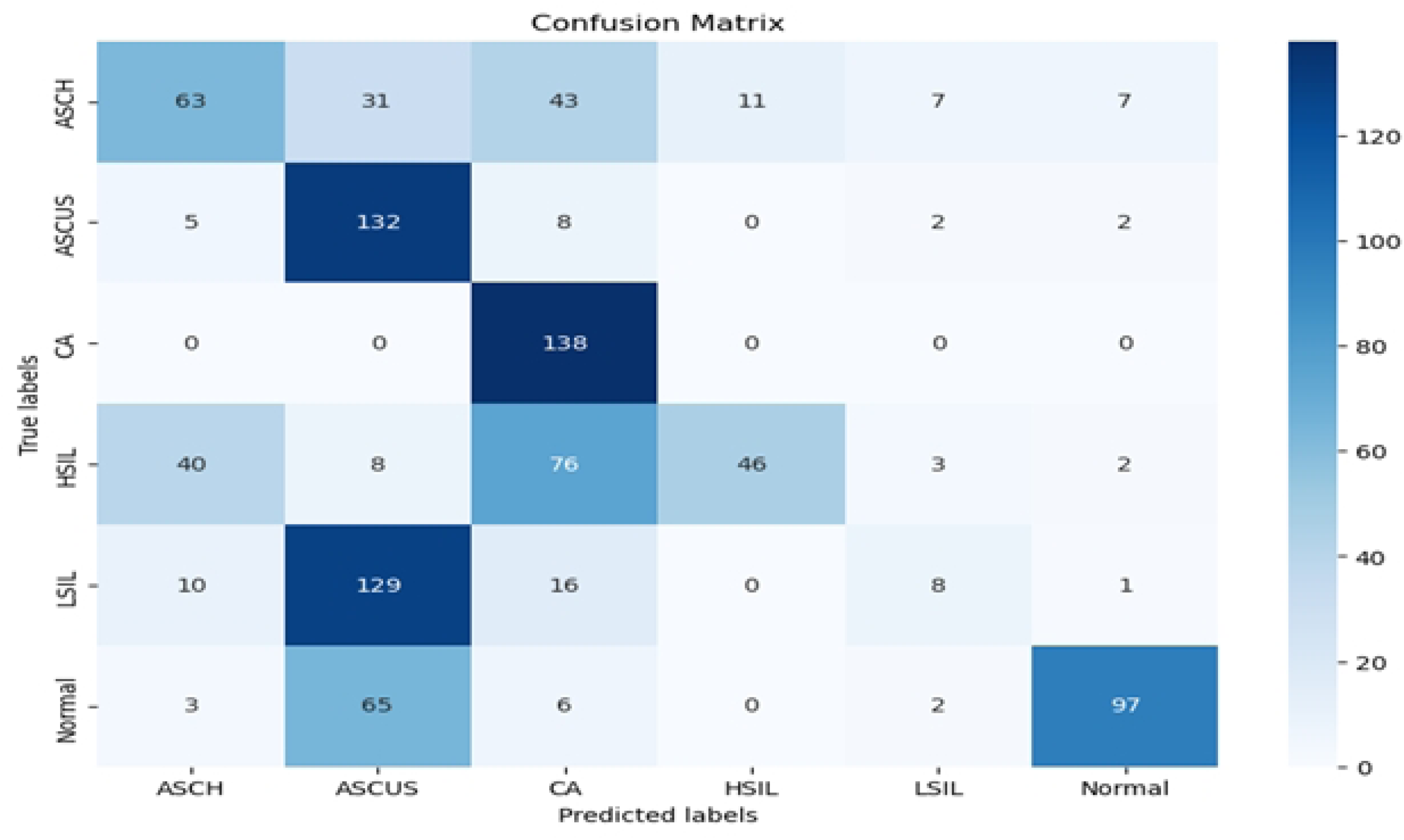
Confusion matrix showing generated EfficientNetB7 performance.

## Discussion

The analysis of the cervical cell dataset from the CRIC Searchable Image Database represents a pivotal step forward in automating cervical cell classification using deep learning (DL) models [14]. By leveraging the CRIC dataset, this study evaluated the performance of various convolutional neural network (CNN) architectures, including EfficientNetB7, MobileNet, ResNet50, ResNet152, and InceptionNet-V3, against a comprehensive spectrum of cervical cell abnormalities as per the Bethesda System [15]. Among these, EfficientNetB7 demonstrated superior performance, indicating its high accuracy in classifying cervical cells into six predefined categories [16]. This finding significantly contributes to the ongoing efforts to improve clinical diagnostics and patient management through AI, addressing the knowledge gap regarding the efficacy of different CNN architectures in handling the complex nature of cervical cell images [17].

The study’s findings highlight the potential of EfficientNetB7 in enhancing the precision of cervical cell classification, a crucial aspect of cervical cancer screening and diagnosis [18]. The superior performance of EfficientNetB7, as evidenced by its high scores across several metrics, aligns with the existing literature that acknowledges the architecture’s efficiency and accuracy in medical image analysis [19]. This contrasts with the variable effectiveness of other tested CNN architectures, notably InceptionNet-V3, underscoring the importance of model selection based on the dataset’s specific characteristics [20]. The challenges identified in distinguishing between certain cervical lesion classes, such as ASC-US and LSIL, reflect the nuanced nature of cytomorphological criteria, resonating with discussions in the literature about the limitations of image-based analysis in capturing detailed cellular features [21]. These insights contribute to a broader understanding of the potential and limitations of applying AI in cervical cancer screening, emphasizing the need for model optimization and methodological advancements to address classification challenges [22].

One of the study’s limitations is the use of a dataset collected from the global North, which may not fully represent the diversity of cervical cell abnormalities present in the global South [23]. This geographic bias highlights the need for future research to include more diverse datasets that better reflect the global incidence of cervical cell abnormalities [24]. Additionally, the difficulty in distinguishing between certain lesion types suggests a potential direction for future research to explore whole smear analyses or the integration of additional contextual information to improve classification accuracy [25].

Despite these limitations, the study’s findings underscore the critical role of AI in advancing cervical cancer diagnostics [26]. EfficientNetB7’s effectiveness suggests that AI can significantly contribute to the accurate classification of cervical cells, thus enhancing patient management strategies [10]. Future research should focus on overcoming the identified limitations by incorporating more diverse datasets and exploring innovative analytical methods [27]. Policy-related recommendations may include encouraging the development of AI-based diagnostic tools that are tailored to the specific needs of different regions, particularly those with high burdens of cervical cancer, to reduce global health disparities [28].

## Conclusion

This study marks a significant advancement in the field of cervical cancer diagnostics by leveraging deep learning models to automate the classification of cervical cells, with EfficientNetB7 emerging as the most accurate CNN architecture for this purpose. Its ability to precisely classify a wide range of cervical cell abnormalities heralds a new era in clinical diagnostics and patient care, offering a more efficient and reliable method for screening and diagnosing cervical cancer. The research also underscores the challenges of model selection and the limitations of current datasets, emphasizing the need for further advancements in AI methodology and the inclusion of more diverse data to ensure the global applicability of these tools. Addressing these issues is crucial for fully realizing the potential of AI in reducing health disparities and enhancing cervical cancer management strategies worldwide. Recommendations for future work include a focus on diversifying datasets and exploring new analytical approaches to improve the accuracy and applicability of AI-based diagnostic tools across different populations.

## Data Availability

All data used in this study are publicly available at https://database.cric.com.br/

## Acknowledgment

The authors would like to express their sincere gratitude to Muhimbili University of Health and Allied Sciences (MUHAS) and Muhimbili National Hospital for providing a conducive environment for research and clinical collaboration. We also extend our appreciation to the Center for Recognition and Inspection of Cells (CRIC) for the dataset. Special thanks go to Ocean Road Cancer Institute for its contributions to oncology research and patient care. Lastly, we acknowledge the dedicated efforts of the SarataniAI team for their commitment to advancing AI-driven solutions in healthcare.

